# Development, content validation and reliability assessment of the Bowel Irrigation Questionnaire, a Patient-Reported Experience Measure for Transanal Irrigation: a multi-methods study

**DOI:** 10.64898/2026.07.16.26358225

**Authors:** Emily Farrow, Rogini Balachandran, Rebecca Embleton, Klaus Krogh, Paul F Vollebregt, Julie Cornish, Peter Christensen

**Author notes:** Corresponding Author: Emily Farrow.

## Abstract

**Objective:** To develop the Bowel Irrigation Questionnaire (BIQ), a patient-reported experience measure (PREM) designed to assess the users’ experience of transanal irrigation (TAI) and conduct preliminary evaluation of its content validity and temporal stability.

**Design:** A four-stage multi-methods study comprising: (1) item generation through literature review and qualitative interviews; (2) international three-round Delphi survey and consensus meeting; (3) content validity assessment through cognitive interviews and the Questionnaire on Questionnaires (QQ-10); (4) temporal stability assessment.

**Settings:** Item generation and the Delphi survey were conducted online with healthcare professionals (HCPs) and TAI users from the UK, Denmark, Sweden, Ireland and the Netherlands. Participants in content validity and temporal stability assessment were recruited from a UK site.

**Participants:** HCPs and TAI users were involved throughout development: (1) nine completed qualitative interviews (6 HCPs, 3 TAI users; 4 male, 5 female); (2) 20 completed the Delphi survey (12 HCPs, 8 TAI users; 7 male, 13 female) and (3) six TAI users completed cognitive interviews. Thirty-two TAI users (30 female, 93.8%; median age 52 years, IQR 34-54) completed the BIQ, followed by QQ-10 and repeated BIQ.

**Results:** Multi-disciplinary qualitative interviews and literature review generated 215 statements across 20 sub-domains, refined to 73 statements for Delphi survey. Following Delphi survey and cognitive interviews, the 16-item BIQ addressed training and support (one item); usability (five items), and user experience (10 items). Median QQ-10 value and burden scores were 21 (IQR 19-22.5) and 0 (IQR 0-2.3) respectively, supporting acceptability and face validity. Test-retest exact agreement ranged from 83.9% to 100% across items, with 100% agreement within one response category for every item.

**Conclusions:** The Bowel Irrigation Questionnaire is a PREM assessing the user experience of TAI. In preliminary assessment, the BIQ demonstrates good measurement properties; however, further psychometric evaluation is required before it can be recommended in clinical practice.

**Strengths and Limitations:**

- The item generation process was thorough and well-grounded in both literature review and qualitative interview, ensuring strong content validity from the outset.
- Transanal irrigation users were actively involved at every stage of development, supporting its relevance and use in real-world settings.
- Using an online Delphi survey meant we benefited from the input of topic experts from a range of countries whilst maintaining anonymity to minimise the influence of dominant individuals.
- This study represents the initial stages of PREM development; a larger study in a broader patient population is required to fully evaluate the psychometric properties of the instrument, including construct validity and internal consistency.
- No a priori content validity index thresholds or predefined criteria for removing sub-domains were specified; instead, decisions were made iteratively based on quantitative rating and qualitative feedback to balance content validity and patient burden. This may have influenced the final composition of the instrument.

## Introduction

Transanal Irrigation (TAI) is used to treat a range of conditions, including faecal incontinence, chronic constipation, neurogenic bowel dysfunction and low anterior resection syndrome (LARS). Evidence from randomised controlled trials and observational studies supports its clinical effectiveness (1)(2)(3)(4)(5)(6). TAI for faecal incontinence and constipation was incorporated into NICE guidelines in 2018 (7). Widely used pyramid models place TAI as a treatment escalation in patients whose symptoms persist despite conservative treatment, prior to considering surgical management (8)(9)(10). Expert guidance emphasises supervised training and structured follow-up, with the choice of device and irrigation regimen tailored to the individual (11)(12)(13).

TAI use is increasing in England, with the number of patients receiving community-dispensed prescriptions rising by 72% between 2020 and 2025 (14). Despite this uptake and a variety of available systems, studies demonstrate a reluctance to initiate treatment and limited adherence at 12 months (8)(9)(10). It is hoped that a validated patient-reported experience measure (PREM) will aid healthcare professionals (HCPs) to systematically evaluate modifiable aspects of treatment and comparisons between systems, with the potential to improve adherence and support the development of new TAI devices.

The expanding range of TAI products makes device selection challenging for HCPs, who must consider patients’ ability to use the device and their underlying condition. Designing and selecting devices for home use is further complicated by users’ diverse ages, abilities and home environments. Systematic reviews highlight gaps in collecting user feedback on home medical devices (11)(12). Furthermore, it has been demonstrated that many usability challenges only emerge post-market (13). Although decision algorithms can support device selection (14)(15)(16), a PREM specifically designed to assess the user experience of TAI could further inform clinical decision-making and identify patients who may benefit from device modification or change.

As TAI is used to treat a broad range of conditions, treatment success is often measured using condition-specific patient-reported outcome measures (PROMs), such as the Cleveland Clinic Constipation score, St. Mark’s incontinence score and the LARS score (17)(18)(19), or generic quality-of-life PROMs, such as the Short Form-36 (20)(21). While these instruments assess condition severity or general health status, they do not capture the user’s experience of TAI. We define TAI user experience as user’s perceptions of performing irrigation, including the training and support, practical and physical aspects of device use and integration into daily life. No PROM or PREM currently exists to compare the user experience of TAI across different conditions or devices. The development and validation of a TAI-specific PREM has therefore been identified as a key research priority (6).

With active involvement from international expert clinicians and TAI users throughout, we aim to develop a PREM assessing adult’s experience of TAI. The BIQ is intended to identify areas for improvement, support discussions between users and HCPs and inform service evaluation, rather than assess bowel symptom severity or treatment effectiveness. This study represents the first phase of validation, with potential future applications in decision-making, product comparison and TAI research.

## Methods

Development and early validation of the Bowel Irrigation Questionnaire (BIQ) consisted of four stages:

(1) Statement generation from literature review and qualitative interviews

(2) International online Delphi survey to identify the most relevant statements for measuring the construct

(3) Face and content validity was evaluated through Questionnaire on Questionnaires (QQ-10) and cognitive interviews

(4) Temporal stability was evaluated by administering the BIQ to product-users on two separate occasions

This study was guided by the ACCORD (Accurate Consensus Reporting Document) and COSMIN (Consensus-based Standards for the selection of health Measurement Instruments) guidelines (22)(23).

### Stage One: Item Generation

Nine 60-minute qualitative interviews were conducted to explore the experiences of HCPs and TAI users. Tailored interview guides for HCPs and TAI users were developed based on literature review and market research (Appendix 1,2). Interviews were conducted by a qualitative researcher (AS) and observed by two clinicians (EF TH). Two reviewers (RB TH) independently analysed transcripts to generate statements. Their findings were compared, and differences in statement wording were resolved through discussion between reviewers. The research team reviewed the resulting 215 statements, removing duplication and refining wording for clarity.

### Stage Two: Online Delphi Survey

A Delphi consensus exercise was undertaken to integrate findings from the literature review and qualitative interviews with the perspectives of TAI users and HCPs. This was led by RB and EF who collectively bought industry and clinical experience in TAI. The online format facilitated international participation, while anonymous rating aimed to minimise the influence of professional hierarchy. The Delphi consensus exercise was not prospectively registered.

TAI users aged ≥18 years or older who could independently complete an English-language online survey were eligible. TAI users were eligible after at least 3 months use, enabling inclusion of both relatively new users and long-term users. Eligible HCPs included specialist nurses, gastroenterologists and colorectal surgeons with experience initiating and troubleshooting TAI; all but one HCP had more than 10 years’ experience. No numerical recruitment target was prespecified, recruitment prioritised relevant clinical expertise and lived experience. HCPs were recruited through direct outreach to relevant experts, professional referrals and existing research collaborators. The Delphi survey was pilot tested by members of the research team before distribution to participants to assess clarity of instructions, technical functionality and completion time.

A three-round online reactive Delphi survey was conducted, with participants rating items generated from Stage One. In Round One (December 2024), experts rated their agreement that each item reflected a positive user experience on a 6-point agreement scale and could provide optional free-text comments. Experts were given 10 days to complete the survey, with a reminder sent. Items reaching consensus for inclusion or exclusion were removed, while remaining items progressed to subsequent rounds. Participants received an anonymised summary of the responses from the first round and were invited to participate in the second round of the survey in January 2025 and a third round in February 2025.

### Data Analysis and Consensus

Delphi survey data were analysed using Microsoft Excel. Expert responses to item-level agreement scales were coded into “Top 2 Box” score (T2B) (very high/ high agree), “Medium 2 Box” score (M2B) (somewhat agree/ disagree) and a “Bottom 2 Box” score (BTB) (very high/ high disagree). An item-level content validity index score was then calculated for each item as the number of experts selecting T2B relative to the total number of experts.

Items achieving ≥85% agreement overall and ≥80% within each expert cohort were included in the final BIQ, while those achieving ≤40% overall and ≤45% agreement in each cohort were excluded from subsequent voting rounds (Table 1). Free-text comments were reviewed by the research team; no formal qualitative analysis was undertaken. This process was repeated across three rounds, with minor adjustments to the inclusion and exclusion thresholds to support development of a concise and clinically practical instrument (Table 1).

**Table 1.** Item-content validity index criteria for statement inclusion and exclusion during Delphi process. ^1^Each stakeholder group refers to users, nurses and physicians separately.

| Decision | Panel | Round 1 | Round 2 | Round 3 |
|---|---|---|---|---|
| Inclusion | Overall Panel | ≥85% | ≥85% | ≥85% |
| Inclusion | Each stakeholder group<sup>1</sup> | ≥80% | ≥80% | ≥80% |
| Exclusion | Overall panel | ≤40% | ≤55% | ≤55% |
| Exclusion | Each stakeholder group<sup>1</sup> | ≤45% | ≤65% | ≤65% |
| Additional Inclusion Criterion | TAI users | - | - | 100% |
| Additional Exclusion Criterion | TAI users | - | - | ≤45% |

### Stage Three: Content Validity Assessment

Virtual cognitive interviews were conducted by a clinician (EF), with previous experience of cognitive interviewing, using a semi-structured interview guide (Appendix 3). Using the verbal probing technique, TAI users explained their responses and commented on item interpretation, wording, recall period and response options. Interviews were conducted in two rounds of three participants, with iterative refinement until no major comprehension issues remained.

The QQ-10 was mailed to 35 TAI users, with 32 TAI users completed the BIQ, followed by the QQ-10 and repeated BIQ one to six weeks later for reliability assessment. Established TAI users were recruited from a single site (County Durham and Darlington NHS Trust). Five-point Likert (strongly disagree to strongly agree) were converted to a 0-4 scale. Scores for “value” (items 1-6; maximum 24) and “burden (items 7-10; maximum 16) were calculated by summing item scores. Higher value scores and lower burden scores indicate greater face validity.

### Stage Four: Temporal Stability Assessment

The temporal stability of the BIQ was assessed using a test-rest design. Paper-based versions of BIQ and QQ-10 were posted to participants for completion, with a reminder sent after 4 weeks. Participants were asked to complete the BIQ on two occasions, one week apart. This interval was chosen to minimise recall of previous responses while reducing the likelihood of meaningful changes in participant’s experiences of TAI. The QQ-10 was completed at the initial administration only.

For item-level test-retest reliability, the sample size was based on detecting a linearly weighted kappa of 0.80 against a minimally acceptable value of 0.50, with 80% power (31). For a four-category response scale, the estimated requirement was 31 participants. Test-retest agreement was evaluated using exact agreement, agreement within one response category, Spearman’s rank correlation coefficient and weighted kappa coefficients. Weighted kappa values were interpreted according to established criteria, with values above 0.80 indicating excellent agreement (32).

### Patient and Public Involvement

Patients were not involved in designing the study. Individuals with experience of transanal irrigation (TAI) contributed to questionnaire development through qualitative interviews and as expert panel members in the Delphi survey, ensuring that the questionnaire reflected issues important to TAI users. Users also participated in cognitive interviews and questionnaire evaluation. took part in cognitive interviews to assess clarity and acceptability and completed face validity and test-retest reliability assessments. Study participants will be offered a plain-language summary of the findings and notified when the study is published.

## Results

An overview of the four-stage process used to develop and validate BIQ is summarised in Figure 1.

**Figure 1.**
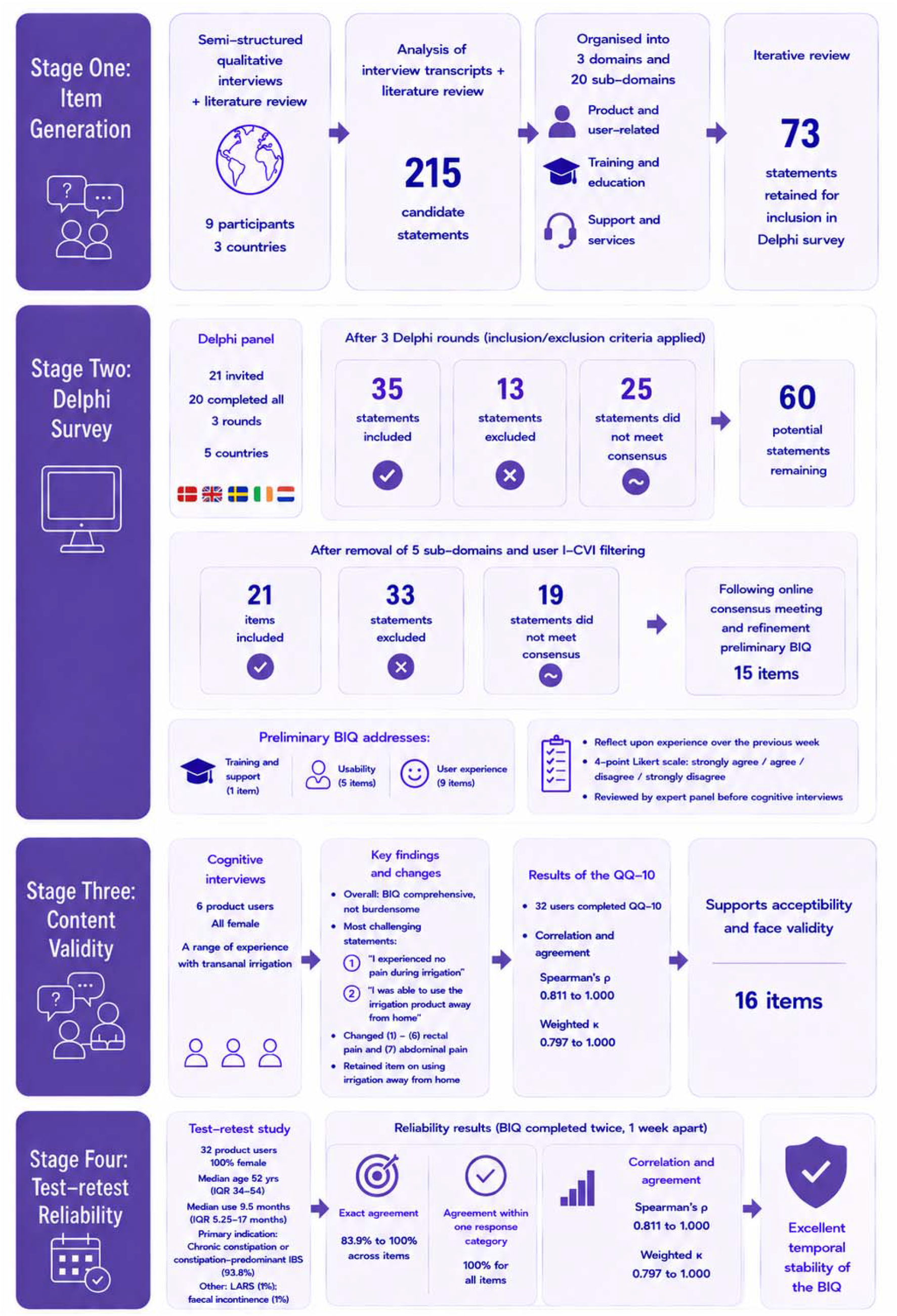
Development and validation of the Bowel Irrigation Questionnaire (BIQ) TAI(transanal irrigation); I-CVI (item-level content validity index); QQ-10 (questionnaire of questionnaires); IQR (interquartile range); LARS (Low Anterior Resection Syndrome)

### Stage One: Item Generation

Nine participants, representing a range of professional and user perspectives on TAI, participated in semi-structured qualitative interviews. The nine participants originated from five different countries (UK, Denmark, Sweden, Ireland and the Netherlands.), five (55.6%) were female and three (33.3%) were TAI users. TAI users’ diagnoses included rectocele, chronic constipation and neurogenic bowel dysfunction. Their experience of TAI ranged from 6 months to over 5 years. Analysis of these interview transcripts, supplemented by literature review, generated 215 candidate statements. These were divided into three over-arching domains: (1) product and user-related, (2) training and education, and (3) support and services, comprising 20 sub-domains (Table 2). Following iterative review, 73 statements were retained for inclusion in the Delphi survey.

**Table 2.**
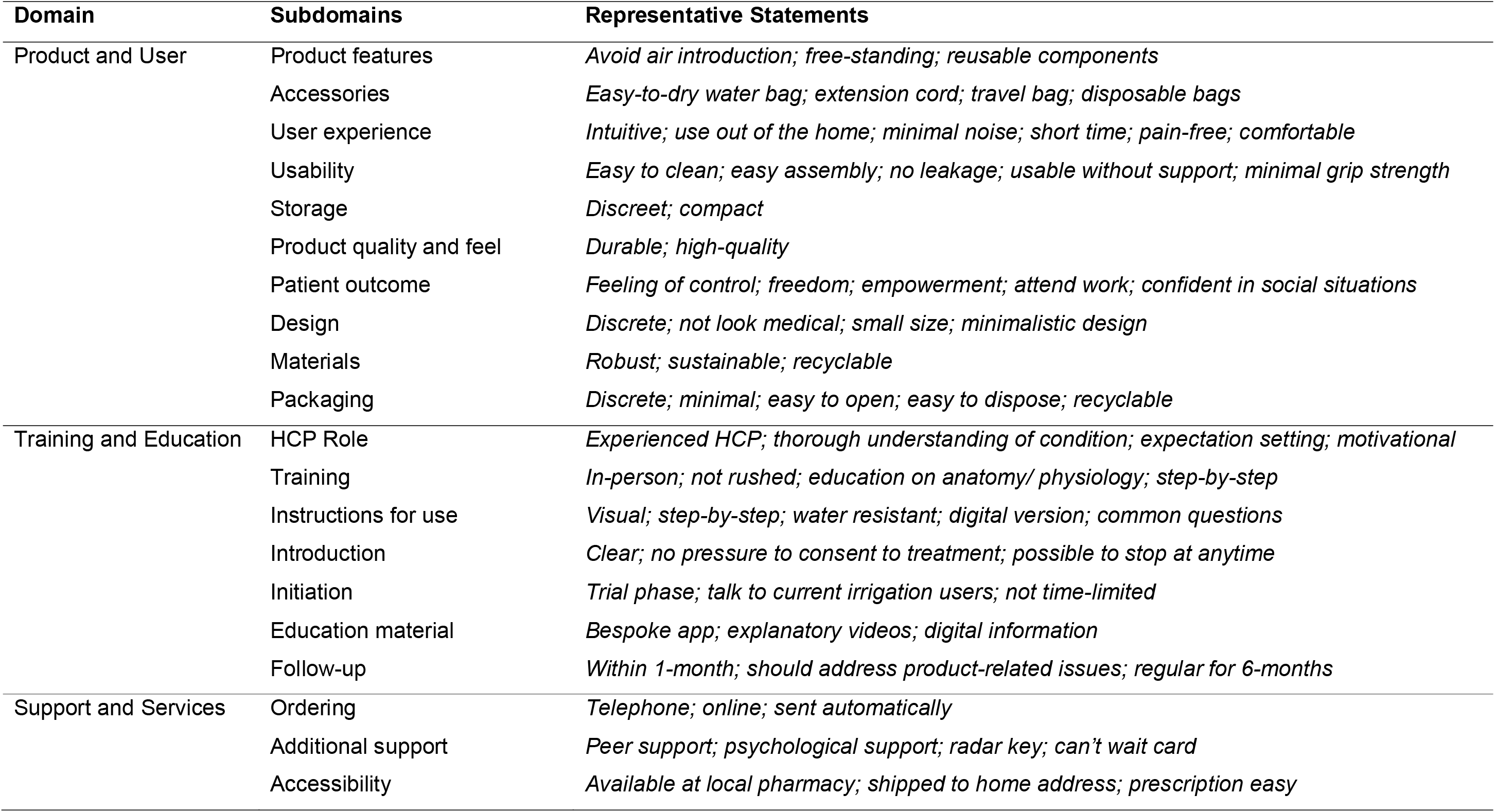
Domains, subdomains and representative statements generated from qualitative interviews.

### Stage Two: Online Delphi Survey

Of the 21 individuals sent the survey, 20 responded to all three rounds. These 20 participants originated from five different countries (Denmark, UK, Sweden, Ireland and the Netherlands), 13 (65%) were female. Twelve were HCPs; seven specialist nurses and five physicians specialising in gastroenterology or colorectal surgery, all with experience initiating TAI across diverse diagnoses. Eight (40.0%) were TAI users with faecal incontinence, chronic constipation and neurogenic bowel and between 3 months and over 5 years’ TAI experience.

After applying the inclusion and exclusion criteria (Table 1) across three rounds, 35 statements were directly included, 13 were excluded, and 25 did not meet consensus. This left 60 potential statements following three rounds of the Delphi survey. Five sub-domains (instructions for use, ordering, accessibility, additional support, and follow-up) were subsequently removed based on expert rankings, eliminating 15 statements. Further exclusion criteria based solely on user opinion were applied, with items achieving a user I-CVI of 100% retained (2 statements) and those with a user I-CVI below 45% excluded (5 statements). This process left 40 items, 19 of which had not yet reached consensus. Following an online consensus meeting, led by RB, and refinement with the research group (RB EF), 29 items remained. Similar items were then reworded and combined, resulting in a preliminary 15 item BIQ, ordered to reflect the typical patient journey through the TAI process. Changes and reasons for retention, merging or exclusion were documented in an item-tracking table (Appendix 4).

The preliminary 15 item BIQ addressed training and support (1 item); usability (5 items), and user experience (9 items). Each of these may be prioritised depending on who is administering the BIQ, for example a clinician may prioritise user experience whereas product developers may prioritise usability. Following item generation and refinement, recall period and response options were developed for the BIQ. TAI users were asked to reflect upon the previous week and rate their agreement with statements on a 4-point Likert scale (strongly agree/agree/disagree/ strongly disagree).

### Stage Three: Content Validity Assessment

Six female TAI users, with between 3 months and 9 years’ experience using a range of TAI systems, completed cognitive interviews. Overall, participants considered the BIQ comprehensive without being burdensome. Minor revisions were made to item wording, format and chronology following participant feedback. A one-week recall period was considered appropriate for most items; however, responses to the training and support item consistently reflected participant’s initial clinician-led training session, irrespective of how long they had been using TAI, resulting in formatting changes.

The two statements participants found the comprehension of most challenging were (1) ‘I experienced no pain during irrigation” and (2) “I was able to use the irrigation product away from home”. Users tended to focus on anorectal pain and did not consider abdominal pain; if they had experienced one or the other, they were then uncertain how to answer the statement. This statement was subsequently divided into (6) rectal pain and (7) abdominal pain, increasing the BIQ to 16 items. Participants generally interpreted “away from home” as another private residence, rather than public toilet, which they considered unrealistic. Despite this, the question was retained in its original form to ensure the BIQ could capture experiences in all environments outside the user’s home.

Thirty-two TAI users completed the QQ-10 following completion of the BIQ, of these 30 (93.8%) were female. Quantitative assessment demonstrated positive feedback (Figure 2,3). There were high median scores for the “value” domains (median of 21 (IQR 19-22.5)). There were low median scores for the “burden” domain (median of 0 (IQR 0-2.3)), suggesting acceptability and face validity of the tool.

**Figure 2.**
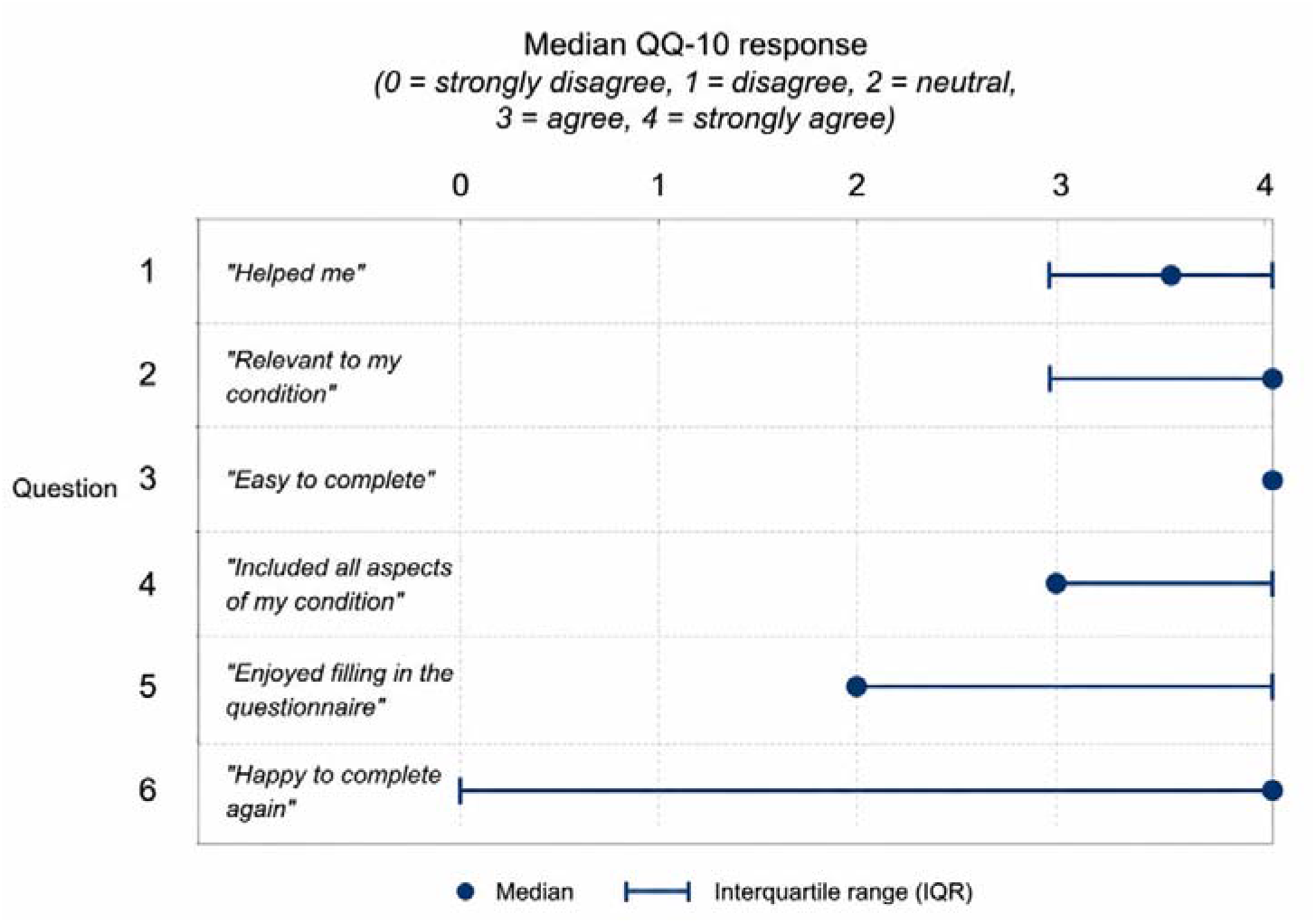
Distribution of QQ-10 Items ("Value") Questions 1-6: Median and Interquartile Range.

**Figure 3.**
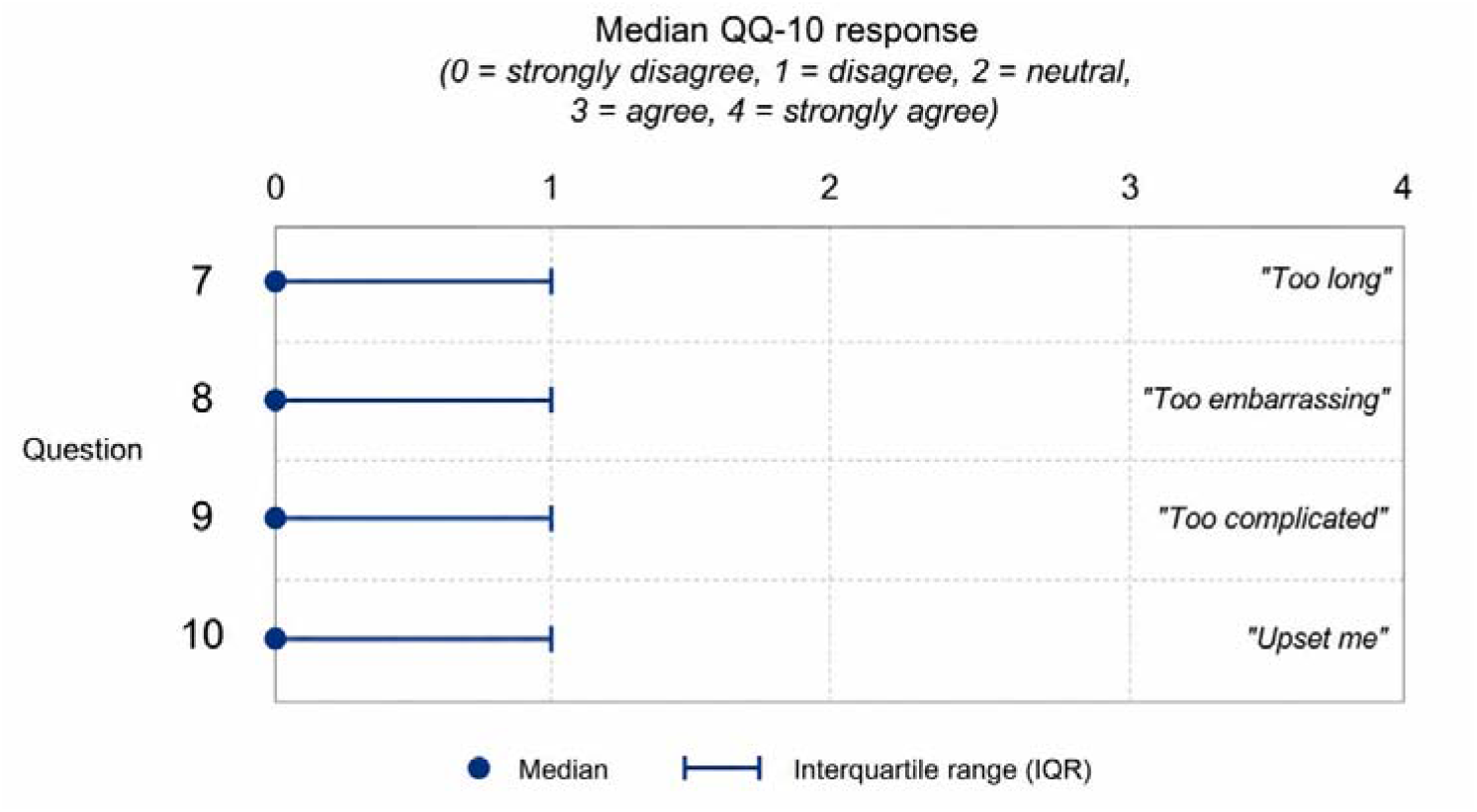
Distribution of QQ-10 Items ("Burden") Questions 7-10: Median and Interquartile Range.

### Stage Four: Temporal Stability Assessment

The same thirty-two TAI users completed the BIQ on two occasions between one and six weeks apart. Participants were predominantly female (93.8%), with a median age of 52 years (IQR 34-54 years). The primary indication for TAI was chronic constipation or constipation-predominant irritable bowel syndrome (93.8%). Participants had been using TAI for a median of 9.5 months (IQR 5-17 months). Established users with longstanding conditions were selected on the assumption that their clinical status would remain relatively stable over the assessment period. Test-retest analyses were conducted using paired responses for each item. There were 16 missing response items, out of a total of 512, equating to 3.1% missing data. Missing responses were not included, test-retest analyses were conducted using all available paired observations for each item.

The results indicate a strong stability with exact agreement ranging from 83.9% to 100% across items, while agreement within one response category was 100% for all items (Table 3). Spearman’s rank correlation coefficients ranged from 0.811 to 1.000, and weighted kappa coefficients ranged from 0.797 to 1.000, demonstrating strong consistency over time. The lowest reliability estimates were for Q6 and 7 (“I was free of abdominal” / “I was free of rectal pain during irrigation”), κ= 0.840 and κ=0.797 respectively. Overall, these findings provide preliminary evidence of generally high test-retest reliability for the BIQ. However, the estimates should be interpreted cautiously because clinical stability between administrations was not formally verified.

**Table 3.** Test-retest agreement for individual BIQ items.

| Item | Exact agreement, % (n/N) | Agreement within one category % (n/N) | Spearman’s ρ | Weighted κ |
|---|---|---|---|---|
| *1* | 100.0% (32/32) | 100% (32/32) | 1.00 | 1.00 |
| *2* | 96.9% (31/32) | 100% (32/32) | 0.82 | 0.88 |
| *3* | 96.9% (31/32) | 100% (32/32) | 0.99 | 0.96 |
| *4* | 93.8% (30/32) | 100% (32/32) | 0.95 | 0.91 |
| *5* | 93.5% (29/31) | 100% (31/31) | 0.91 | 0.91 |
| *6* | 86.7% (26/30) | 100% (30/30) | 0.87 | 0.84 |
| *7* | 83.9% (26/31) | 100% (31/31) | 0.81 | 0.80 |
| *8* | 96.9% (31/32) | 100% (32/32) | 0.92 | 0.91 |
| *9* | 96.9% (31/32) | 100% (32/32) | 0.91 | 0.90 |
| *10* | 96.9% (31/32) | 100% (32/32) | 0.92 | 0.93 |
| *11* | 96.9% (31/32) | 100% (32/32) | 0.96 | 0.96 |
| *12* | 96.9% (31/32) | 100% (32/32) | 0.93 | 0.94 |
| *13* | 96.7% (29/30) | 100% (30/30) | 0.96 | 0.96 |
| *14* | 90.0% (27/30) | 100% (30/30) | 0.93 | 0.89 |
| *15* | 93.5% (29/31) | 100% (31/31) | 0.98 | 0.93 |
| *16* | 96.3% (26/27) | 100% (27/27) | 0.86 | 0.88 |

## Discussion

The Bowel Irrigation Questionnaire (BIQ) is a novel PREM designed to assess the user experience of TAI (Figure 4). Expert clinicians and TAI users were involved throughout development, with priority given to the user perspective. With an increasing number of TAI devices available, a standardised assessment of user experience is needed to enable meaningful comparison between devices and support evidence-based device selection. Although created in collaboration with Qufora (Denmark), the BIQ is designed to evaluate user experience across all TAI devices and is not specific to any individual manufacturer or product.

**Figure 4.**
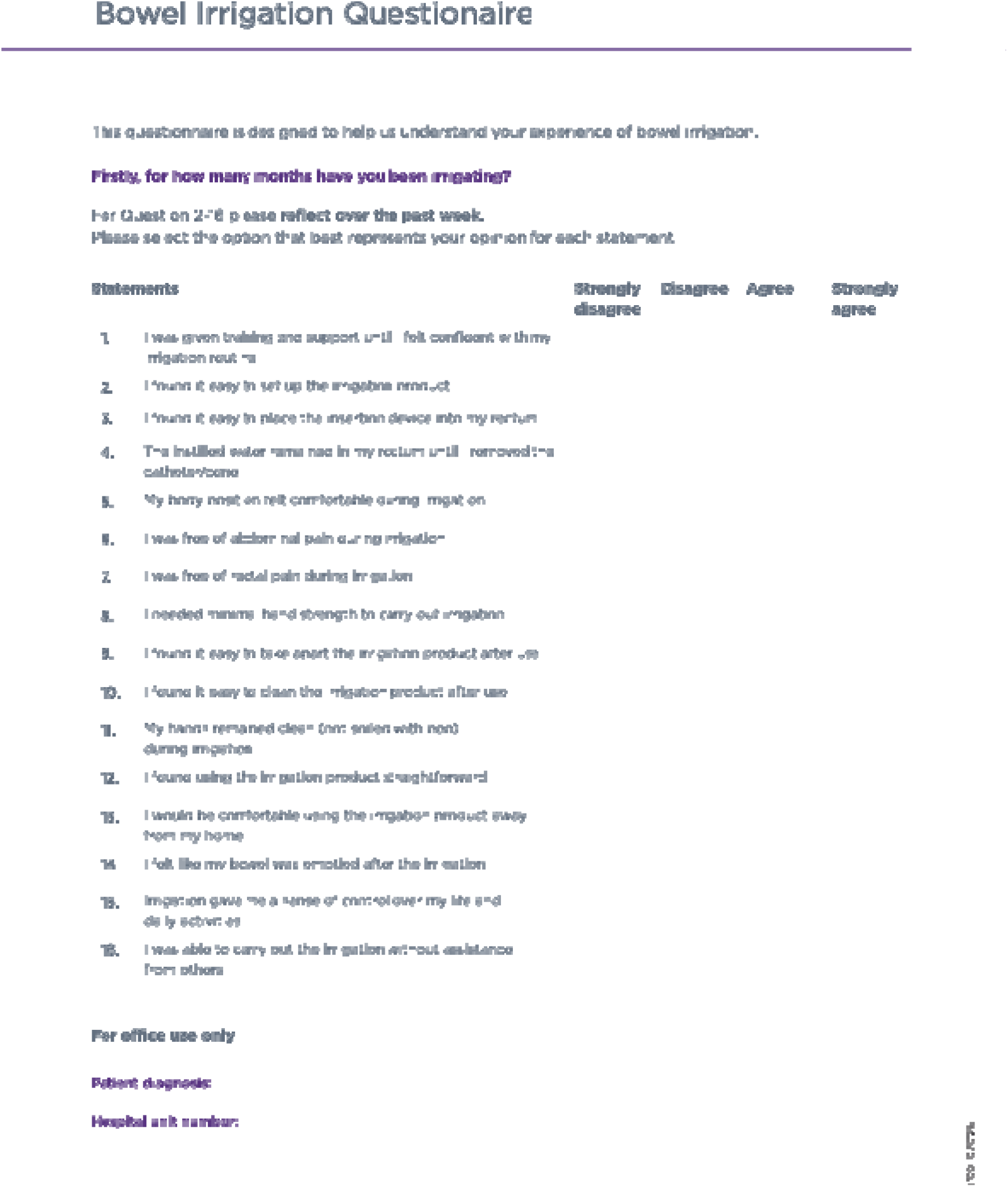
The finalised version of Bowel Irrigation Questionnaire containing 16 items.

PROMS typically assess symptoms and quality of life, whereas PREMs assess the experience of care. Although PROMs and PREMs are traditionally captured separately, patients’ experience and outcomes are interconnected (34)(35)(36). The BIQ reflects this relationship, with 12 statements addressing user experience and four addressing outcomes: “I felt like my bowel was emptied after irrigation”, “irrigation gave me a sense of control over my life and daily activities”, “I was free of abdominal pain” and “I was free of rectal pain”.

During cognitive interviews, TAI users did not differentiate between experience and outcome questions and valued the opportunity to describe TAI’s impact on their quality-of-life; both contribute to a comprehensive assessment of the user experience of TAI. The BIQ has been classified as a PREM because most items emphasise experience. Used alongside condition specific PROMs, the BIQ could help distinguish inadequate symptom improvement from difficulties with training or device use, informing more targeted discussion. The BIQ therefore complements rather than replaces existing PROMs.

At present the BIQ presents an item-level clinical profile organised into three conceptual sub-domains; no total or sub-domain scores are calculated. Individual responses identify strengths and difficulties to support discussion and troubleshooting between HCP and users. Further research is required to determine whether summary scores are appropriate and to establish their interpretation.

There are some limitations to consider. No a priori content validity index thresholds or predefined criteria for removing sub-domains were specified; instead, decisions were made iteratively based on quantitative rating and qualitative feedback to balance content validity and patient burden. This may have introduced subjectivity and influenced the final composition of the instrument. Delphi consensus has no universal definition; a systematic review found a median agreement threshold of 75%, with only 48.8% of studies setting a threshold a priori (33). High levels of agreement were observed throughout the Delphi process (median I-CVI was 0.75, 0.80 and 0.75 for Rounds 1-3), possibly reflecting the crossover of experts used in both the interview process and the Delphi survey. A threshold of I-CVI>0.85 was applied to limit questionnaire length. Following the final Delphi round, user consensus was given additional weight, with statements achieving 100% agreement among TAI users retained to prioritise user perspective.

The generalisability of findings is limited by the study population, which consisted predominantly of female patients with constipation using Qufora TAI equipment. The findings may not extend to males, other diagnoses or users of other systems.

Sociocultural factors, including attitudes towards intimate procedures, privacy and access to facilities, may influence TAI acceptance and willingness to report difficulties. Although the Delphi panel represented three European countries, subsequent BIQ evaluation was undertaken at a single UK centre and does not establish cross-cultural or linguistic validity. Further validation in more diverse populations and settings is warranted.

## Conclusion

We present the Bowel Irrigation Questionnaire (BIQ), a tool consisting of 16 items rated using a 4-point Likert scale, designed to assess the user experience of TAI. The BIQ was developed through a structured process of item generation, refinement and preliminary testing. Preliminary evidence suggests that the instrument demonstrates good content validity and temporal stability in female patients with constipation. Further independent psychometric evaluation in broader patient populations, including men and patients with other diagnoses, and across different TAI products is required before BIQ can be recommended in clinical practice.

## Data Availability

All data produced in the present study are available upon reasonable request to the authors.

## Funding Statement

This study was funded by Qufora. Health care professionals and product user participating in the qualitative interviews and Delphi survey received reimbursement for their time. Reimbursement was provided solely for participation in study activities and was not linked to authorship or study outcomes. Data analysis and interpretation were conducted independently by the study team.

## Conflict of Interests

One author (RB) is an employee of the funder and contributed to the study design and coordination. Some health care professionals who participated in the expert Delphi panel (KK, PV, JC, PC) were also study authors and contributed to the study design.

Paul F. Vollebregt and Klaus Krogh have received honoraria from Coloplast A/S and Qufora for serving on advisory boards and/or working as a speaker. Julie Cornish has received an educational grant from BD & Medtronic and is Clinical Director of Everywoman Festival. Peter Christensen received honoraria from Coloplast A/S for consultancy, teaching, and as an Advisory Board member, and from Wellspect and Qufora. All other authors declare no conflicts of interest.

## Ethics

Ethical approval was not required for this study following consultation with the Health Research Authority (HRA) Research Ethics Committee (REC) in Newcastle. The study involved service evaluation of existing product users with no additional burden. All participants provided informed consent prior to participation and data were anonymised for analysis.

## Notes

### Author Declarations

The Health Research Authority (HRA) Research Ethics Committee (REC) in Newcastle waived ethical approval for this work. The study involved service evaluation of existing product users with no additional burden. All participants provided informed consent prior to participation and data were anonymised for analysis.

### Summary of Updates

We have revised the title to reflect the multiple methods used and specify the measurement properties evaluated. The abstract has been restructured in accordance with BMJ Opens Instructions for Authors, with distinct Design, Setting and Participants sections. We have also expanded the abstracts Results section in response to your comments. In response to Reviewers 1 and 2, we have expanded the first two paragraphs of the Introduction to provide further context on transanal irrigation and the scale of the problem. We have incorporated the paper by Bazzochi et al. as suggested by Reviewer 1 and defined the construct of TAI user experience. Further detail regarding the qualitative interviews and Delphi survey methodology has been added in paragraphs 2-4 of the Methods. The Results now also provide more information about the product users who participated in these stages of the study, as per Reviewers 1 and 2 comments. In response to both reviewers comments on the inclusion of patient-reported experience and outcome items, we have expanded the Discussion to explain the relationship between these constructs and their relevance to the BIQ. Within the Discussion we have also added a paragraph clarifying, at this stage, the BIQ should be interpreted as an item-level clinical profile and that further work is required to establish whether summary scores are appropriate and psychometrically supported. Both reviewers highlighted the limited generalisability of the BIQ given that it has been validated in a population which is predominantly women with constipation. In both the abstract, discussion and conclusion we have more explicitly stated that further psychometric evaluation is needed in a broader population. Table 1 has been re-formatted to make the consensus thresholds between each round clearer, as requested by Reviewer 1. We have also added a tracking table (Appendix 4) summarising the reduction from 29 candidate statements generated following the Delphi survey to 16 items, together with a completed ACCORD checklist.

