## Appendix 4 for "Development, content validation and reliability assessment of the Bowel Irrigation Questionnaire, a Patient-Reported Experience Measure for Transanal Irrigation: a multi-methods study"

| **Domain** | **Statement** | **ID** | **Item Refinement** | **Finalised Statements** |
| --- | --- | --- | --- | --- |
| **Product-related items** | The device should avoid introducing air into the bowel | ID01 | Exclude |  |
| **Usability** | The irrigation product should be easy to assemble | ID02 | Include, Rephrase | "I found it easy to set up the irrigation product" |
|  | The cone/catheter should be easy to insert (into the rectum) | ID03 | Include, Rephrase | "I found it easy to place the insertion device into my rectum" |
|  | The irrigation product should be easy to use | ID04 | ID04 ID11 Combined | "I found using the irrigation product straightforward" |
|  | The water pump should be usable with minimal grip strength | ID05 | Include, Rephrase | "I needed minimal hand strength to carry out irrigation" |
|  | The irrigation procedure should not cause any leakage of water | ID06 | Include, Rephrase | "The instilled water remained in my rectum until I removed the catheter/cone" |
|  | The irrigation product should be usable without support from others | ID07 | Include, Rephrase | "I was able to carry out irrigation without assistance from others" |
|  | The irrigation product should be easy to disassemble | ID08 | Include, Rephrase | "I found it easy to take apart the irrigation product after use" |
|  | The water bag/device should be easy to dry | ID09 | ID09 ID10 Combined |  |
|  | The irrigation product should be easy to clean | ID10 | ID09 ID10 Combined | "I found it easy to clean the irrigation product after use" |
| **User-experience** | The use of the irrigation product should be intuitive/easy to understand | ID11 | ID04 ID11 Combined |  |
|  | The irrigation product should be easy to use out of home | ID12 | Include, Rephrase | "I was comfortable using the irrigation product away from my home" |
|  | The irrigation procedure should be completed in short time | ID13 | Excluded, Repetition |  |
|  | The use position/body posture while irrigating should feel comfortable | ID14 | ID14 ID26 Combined | "My body position felt comfortable during irrigation" |
|  | The irrigation procedure should be pain-free | ID15 | Include, *separated into abdominal & rectal pain* | "I felt free of abdominal pain during irrigation" "I felt free of rectal pain during irrigation" |
|  | The irrigation procedure should completely empty my rectum | ID16 | Include, Rephrase | "I felt like my bowel was emptied after the irrigation" |
|  | The irrigation procedure should require minimal cleaning of surroundings after use | ID17 | ID 17 ID 18 Combined | "My hands remained clean during irrigation" |
|  | The product use should limit the risk of getting used irrigation water on hands | ID18 | ID 17 ID 18 Combined |  |
|  | Irrigation should give a feeling of control | ID19 | ID 19 ID 20 Combined | "Irrigation gave me a sense of control over my life and daily activities" |
|  | Irrigation should enable participation in roles/situations that are important to me/users | ID20 | ID 19 ID 20 Combined |  |
| **Involvement of health care professionals (HCP)** | HCPs should be experienced in irrigation procedures | ID21 | Exclude |  |
|  | HCPs should present multiple product options | ID22 | Exclude |  |
|  | HCPs should thoroughly examine my condition/situation before advising on irrigation | ID23 | Exclude |  |
|  | HCP support to develop an irrigation routine should be offered | ID24 | Exclude |  |
| **Training and education** | The training should not feel rushed | ID25 | Exclude |  |
|  | The training should help find the best use position/body position | ID26 | ID14 ID26 Combined |  |
|  | The training should cover a step-by-step guide of the procedure | ID27 | Exclude |  |
|  | There should be a trial phase to consider if irrigation is the right solution | ID28 | Exclude |  |
|  | The training should continue until feeling confident in the irrigation procedure | ID29 | ID29 Rephrase | "I was given training and support until I felt confident with my irrigation routine" |
|  | **Total Statements: 29** |  |  | **Total Statements: 16** |

*Appendix 4. Tracking of candidate items included following the Delphi survey, showing domains, original statements and refinement decisions to generate finalised BIQ items.*
