## Appendix 2 for "Development, content validation and reliability assessment of the Bowel Irrigation Questionnaire, a Patient-Reported Experience Measure for Transanal Irrigation: a multi-methods study"

**Appendix Two: Interview Guide for TAI Users**

**INTERVIEW GUIDE: Users**

**Target group: 9 diverse profiles with experience with rectal irrigation,** covering academia, health care professionals and users.

**Length of interviews:** 60 min.	**Internal Ipsos job reference:** 24-052267

| **Introduction** | **5 min** | to | **5 min** |
| --- | --- | --- | --- |
| **Respondent understanding** | **10 min** | to | **15 min** |
| **A good irrigation experience** | **20 min** | to | **30 min** |
| **Parameters throughout journey** | **25 min** | to | **55 min** |
| **Wrap-up** | **55 min** | to | **60 min** |

**Research description to guide your probing**

Qufora is a fast-growing MedTech company focusing on rectal irrigation products aimed at people suffering from chronic constipation and/or faecal incontinence.

In this research, Qufora wants to develop a validated questionnaire that can measure the irrigation experience of rectal irrigation products. This questionnaire will serve both in new product development, product comparison studies and for medical research.

To build this questionnaire, we first need to uncover the different **aspects that affect the irrigation experience** to uncover the relevant statements that affect irrigation, which is the purpose of these qualitative interviews.

**Key research themes and questions:**

- What support elements influence the irrigation experience positively and negatively?
- What training elements	influence the irrigation experience positively and negatively?
- What product-related elements influence the irrigation experience positively and negatively?

**Notes:**

- The research **approaches irrigation holistically** covering both practical, product-related and mental aspects of rectal irrigation.
- The respondents **cover a range of profiles** covering users, nurses, professors and physicians. They are however all experienced and knowledgeable about irrigation.
- Please **keep in mind the target groups**, adjusting the questions and time spent on the different sections depending on the respondent’s situation.
- The terms transanal irrigation, TAI, rectal irrigation, and bowel irrigation may be used interchangeably. Please **find a common term that resonates with the respondent** and use this throughout the interview.
- Overall, we are pursuing **width in insights**, trying to uncover all the elements that influence an irrigation experience. The relative importance of these elements are interesting, and will be covered in later stages of the research process, but it’s not the main objective.

**INTERVIEW GUIDE**

**1. INTRODUCTION & PRESENTATION	(5 min / 5 min)**

***The purpose of this section is to outline the purpose of the interview and the conversation ‘rules’ and clear out any questions prior to the interview.***

Thank you for taking part in our patient study about rectal irrigation:

- The interviewer briefly presents him/herself and Ipsos Healthcare: *Ipsos is an independent research agency, working on behalf of a medical device company, with no specific interest invested in the output of this research.*

*Before we begin, I will go through some practical information…*

**Framing introduction to the interview ensuring consent:**

- We **comply with the code of procedure** from EphMRA, BHBIA and European GDPR guidelines for **data protection.**
- The content of our conversation will only be used for **analytical purposes**.
- **I am interviewing several people** that have experience with rectal irrigation, as well as health care professionals in different roles working with rectal irrigation. We will work all input into findings and conclusions at an aggregated level. There will be no representation at a personal level.
- The conversation is **confidential outside the project group at Ipsos and their client,** Qufora**,** and your participation will remain anonymous in all outcomes.
- The client, a manufacturer of rectal irrigation products, has the opportunity to **observe the interviews.** We have agreed that they will be able to ask a few questions in the end if there are any points they would like to have elaborated.
- When conducting research within healthcare, we are required to pass on to our client details of **adverse events/product complaints** pertaining to their products that are mentioned during the interview. If this happens, we will need to collect details and report the event, even if you have already done so. We have a procedure for this, and I will let you know if an AE is to be reported. In this case, I will ask you whether you consent to assist providing additional information to the client company’s safety department for their follow up, but you may choose to remain anonymous. This has no impact on the confidentiality and anonymity associated with the interview itself.
- I am **recording this interview** to help me remember and secure the best possible reporting. The recording will not be shared and will automatically be deleted 3 months after the end of this project.
- You are **free to stop the interview at any time** if you do not want to continue.
- With this information in mind, **are you happy to continue?** (ensure consent)

When consent is obtained, the interviewer outlines the conversation ‘rules’:

- The objective of this conversation is to **understand your perceptions and considerations around using rectal irrigation products** [may also be called transanal irrigation, anal irrigation or bowel irrigation], and the training and support offered.
  - What term do you prefer? [moderator to adapt language]
- The conversation will take about 60 minutes, give, or take 5 minutes.
- I have some themes and questions I’d like to cover, but it is an **open conversation**, so please do not hesitate to share on what’s on your mind.
- Keep in mind that **we are interested in your thoughts and experiences.** You know what it means to be in your situation and that is why we have invited you to take part in this research.
- Please be open and honest**.** There are **no right or wrong answers**. People are different and have had different experiences. This diversity is exactly what we are looking for.
- If you need a break during our conversation, please tell me.
- Is there anything I should keep in mind or pay special attention to?
- Do you have **any questions before we begin?**

**2. RESPONDENT UNDERSTANDING	(10 min / 20 min)**

***The purpose of this section is understanding the condition, journey and experiences of the respondent.***

**MODERATOR NOTE:** Allow the respondent to tell their story but be very mindful that we do not spend too much time.

2.1 [3 minutes] **User introduction**

**Moderator introduces him-/herself** to begin with, covering the same aspects as we would like the respondent to cover: Occupation, living situation, family situation etc.

- To begin, can you tell me a bit about yourself and your life?
- How would you **describe your bowel issues**?
  - How severe would you say it is?
  - For how long have you experienced these issues?
- How does your bowel issues **affect you today**?
  2.2 [7 minutes] **Experience with rectal irrigation**

Our focus today is on rectal irrigation.

- What is **the first thing that comes to mind** when you think of rectal irrigation?
- Can you tell me about your **history and experience** with irrigation products?
- I would like to **map out your “journey” with rectal irrigation**. Can you guide me through the important steps from the first time you heard about it up until today?
- Can you tell me a story about a **positive experience** you’ve had with irrigation?
- Can you tell me a story about a **negative experience** you’ve had with irrigation?

**3. UNCOVERING A GOOD IRRIGATION EXPERIENCE	(20 MIN / 25 MIN)**

***The purpose of this section is to understand what makes a good irrigation experience throughout the journey.***

3.1 [10 minutes] **Learnings from current device**
- Which product are you using today and what do you think about it? [Probe]
  - For how long have you been using this product?
- Talk me through your current product. What works well, and what could be improved?
  - What is the best thing about the product?
  - What are the biggest challenges with this product?
  - How does it compare to other products you have tried or heard of?

**MODERATOR NOTE:** Spend some time probing here to uncover elements that influence the overall experience. We are seeking width here to derive all important statements. You can use the below list for probing options to drive the conversation but please aim to uncover what is on the respondent’s mind first. Below is to be seen only as a probing aid if needed.

- **Ease of use:** How easy is the product to set up, use, and clean?
- **Use experience:** Comfort, discomfort, feelings during use, insertion
- **Adjustment:** How easy is it to adjust (e.g. water volume, temperature) etc.
- **Effectiveness:** How effective is the product in emptying the bowel? Does it provide predictable and consistent results? How long does the effect last?
- **Consistency / predictability:** Does the product provide consistent and predictable results?
- **Components:** Are there any difficulties with specific components (e.g., catheter, pump)?
- **Instructions:** Are the instructions clear and useful?
- **Safety:** Does the user feel safe using the product? Are there any concerns about potential complications or side effects?
- **Portability:** How is it to use the product outside of home? (if relevant). How is it to travel with the product?
- **Package:** How is the package experienced? Does it matter?
- **Look/design:** How is the look and design of the product experienced?
- **Discretion:** Does the product and pack offer the desired discretion?
- **Training:** How helpful were the initial training and educational materials?
- **Support:** How are the support functions used? What additional support would be beneficial?
- **Impact on life (physical, emotional and social):** How has rectal irrigation affected physical symptoms like bloating, discomfort, and accidents? How has it impacted self-esteem, confidence, and anxiety related to bowel control? Has it enabled greater participation in social activities, work, and travel?
  3.2 [10 minutes] **The ideal product & experience**

Thank you for telling me about your current product. Now I’d like you to envision that you are the lead for developing a new irrigation product ideal for yourself. How would you build that product?

- What is important to keep in mind when developing the product?
- What do you see as the different areas and aspects to focus on?
- Let’s focus a bit on the use experience. Can you describe a good irrigation experience?
- Let’s focus on the product itself. What constitutes a great product for irrigation?
- Let’s cover the surrounding training, support and services. What’s important to offer here?
- Now try to imagine a poor product and experience. Can you describe it?
- What is important to avoid when building an irrigation device?
  - For the product? The experience? The support?

Now, let’s put it together: Can you describe the ideal irrigation product (and experience)?

- Sentence completion exercises:
  - “Irrigation should be…”
  - "Using rectal irrigation should make me feel…"
  - “A good irrigation experience is…” (let me know all the words that come to mind)
  - “My ideal irrigation product would…”
  - “The ideal irrigation product is definitely NOT…”

**MODERATOR NOTE:** Please apply an extensive probing asking “what else?” and “why” rather one time too many on the aspects raised by the respondent. Take your time to

If the respondent struggles with this exercise, then try to include some sentence completion exercises: “A good irrigation experience is…”, “it’s important that I feel… when irrigating”, "The biggest challenge with rectal irrigation is…,", "My ideal irrigation product would…"	etc.

**4. PARAMETERS THROUGHOUT THE JOURNEY	(25 MIN / 55 MIN)**

**In this section we focus on specific areas of interest that we want to uncover. Where the earlier sections have purposefully been open in nature, this section focus on specific elements.**

I will now try to be a bit more specific and detail-oriented and ask about the important parameters throughout the different phases from first introduction to regular use.

4.1 [10 minutes] **Initiation Phase & Training:**

Looking back at the early phases, when irrigation was first introduced to you. **How did you experience this period? How did you feel? What thoughts and concerns were on your mind?**

- What **expectations** did you have for the irrigation device before using?
- What **concerns or hesitations** did you have before starting with irrigation?
  - How did you overcome these concerns?
- What were your **first reactions** seeing the **packaging**?
  - Zooming in on the packaging. What is important when it comes to the pack look and design?
- What in your opinion is key, for a new user to feel **confident and comfortable** starting to use an irrigation product in these early phases?
  - Did you have any contact to support networks or patient groups? If so, how did you experience this?
- Did you receive any **demonstration** of the product before usage? Can you tell me about it and how you experienced it?
  - What worked well and what could perhaps be better?
  - Imagine you were to demonstrate your product to another person. How would you do it?
  - What’s important to keep in mind when demonstrating the product?
  - Did you do a physical trial of the product with a health care professional? If so, how did you experience it?
    - What are your thoughts on having a physical trial?
- Do you recall your first **assembling and dissembling** of the product? How did you experience this?
  - How would you describe the experience?
  - What worked well and what was perhaps more challenging?
  - What would you like producers to have in mind when designing the assembling and dissembling mechanisms?
- How was your **access to health care professionals** for support in this phase?
  - How did you use them?
  - How and who would you have liked to have access to in this phase? And why?
- Did you have access to any **training material**? If so, how did you use it?
  - What worked well and what did not work so well?
  - Is there any material, you would have liked to have?

4.2 [10 minutes] **The irrigation procedure**

**Let’s take a closer look at the irrigation procedure and experience.**

- How easy or difficult do you find doing an irrigation procedure? What could be better / easier, if anything?
- When, how do you typically perform irrigation and why?
  - How would you describe the ideal setting for irrigation?
  - How does the setting impact the experience?
- How do you feel after an irrigation? How does this feeling impact you?
  - What feeling would you like to have after an irrigation?
  - Sentence completion: “Irrigation should leave me feeling…”
- Have you ever experienced any pain or discomfort? Can you describe what happened and how you felt about it?
- How do you perform the irrigation? What to you is the best position or posture?
  - How would you a describe a good use position / posture?
  - How would you design the usage position if you could choose freely?
- Have you ever experienced any leakage/dripping during or after the procedure?
  - How did you feel about it the first time?
  - How do you feel about it today?
  - How much can be accepted?
- Have you ever experienced any other device-related issues? Which and how did you experience them?
- How and where do you store your irrigation device after use? Why there?
  - How easy / difficult is it to store the device?
  - What it’s important to you in terms of storing when you’re not using the device?

4.3 [5 minutes] **Impact on life**

- How has irrigation **impacted your life** after you started doing it regularly? (socially, professionally, emotionally etc.)
  - Sentence completion: “Irrigation has given me…”
  - Sentence completion: “Irrigation has meant that…”
  - Sentence completion: "Irrigation has caused me to…”
- What, if any, **limitations** do you experience on your life related to using an irrigation device?
  - Are there anything, that you would like to do, that you refrain from due to irrigation? (e.g. travel, social activities, events on specific time slots etc.)
  - Are there any limitations that originate in the device? (e.g. the size, the look, the function etc.)
  - What could be changed to minimize any negative impact on your life?
- What, if anything, might **preventing** you from using your irrigation device? Are there any situations where you postpone the irrigation? Tell me about your thoughts here.
  - What could trigger a postponement? (space, constraints etc.)

**5. WRAP-UP	(5 MIN / 60 MIN)**

**In this section we wrap up the conversation and follow-up on any unanswered questions.**

5.1 [5 minutes] **Understanding key focus areas ONLY IF TIME ALLOWS**

Imagine you are speaking to someone who has been recently introduced to rectal irrigation, same as you. Your friend is feeling apprehensive about using rectal irrigation products and has come to you for advice because they know you have experience with these products.

- What’s the first thing you would say?
- What would you highlight as things to be mindful of?
- What expectations would you set out?
- How would you explain the process of using irrigation? What would you tell them about the comfort level, effectiveness, and overall experience?
- How would you address their concerns? Remember, you can speak freely about both the positives and any challenges you have encountered.
- What other advice would you ensure to provide them?

5.2 [5 minutes] **Wrap-up**

- Here at the end, I would like us to summarize together. Taking our full discussion into consideration, what are, in your opinion, the important **parameters** to which you would judge/evaluate an irrigation experience?
  - If you have to sum up the characteristics of “a good device”, what are the five most important things?
- Is there anything that is on your heart or mind that we have not covered today?
- *Ask questions from client/backroom if any.*
- Do you have any questions for us before we wrap-up?

**Thank and close the interview.**
