## Appendix 3 for "Development, content validation and reliability assessment of the Bowel Irrigation Questionnaire, a Patient-Reported Experience Measure for Transanal Irrigation: a multi-methods study"

**Appendix Three: Interview Guide for Cognitive Interviews**

**Interview Guide for Bowel Irrigation Questionnaire Development**

**Version 1.0 22/07/2025**

**Phase One: Cognitive Debriefings**

Before the Cognitive Debriefing:

- Informed consent will be collected from patients who wish to participate before the cognitive debriefing.
- Eligibility criteria of each patient will be completed on confirmation of consent with the patient.
- Participants will be contacted to arrange a convenient day and time for the cognitive debriefing which will take place online or in-person on an individual basis.
- A clinical researcher will carry out cognitive debriefings. There will also be a researcher with clinical expertise in TAI observing the interview.
- Participants will be sent a copy of the Bowel Irrigation Questionnaire to review at least one week before the focus group.

Eligibility Criteria

Inclusion Criteria:

- Adult patients aged ≥ 18 years
- Able to provide written informed consent
- User of Transanal Irrigation for at least 6 months?
- Can speak English

Start of Cognitive Debriefing:

- Confirm the participant is happy to take part in the cognitive debriefing.
- Advise that they can withdraw consent at any stage and skip over any questions they don’t wish to answer without providing a reason.
- Introductions to all participants.
- Supply with new copies of the questions if needed

**Part One: Comprehension**

1) I was given training until I felt confident with my bowel irrigation routine.

**Comprehension:** What does training mean to you? What does support mean to you? What does confident with your routine mean to you?

**Decision:** How easy or difficult is it to answer this question?

**Response:** How did you arrive at your answer?

2) The irrigation product was easy to set up for use.

**Comprehension:** What does irrigation product mean to you? What does easy to set up mean to you?

**Decision:** How hard is it to answer this question?

**Response:** How did you arrive at your answer?

3) It was easy to place the insertion device into my rectum

**Comprehension:** What does easy to place mean to you? What does insertion device mean to you? Do you understand the term rectum?

**Decision:** How hard is it to answer this question?

**Response:** How did you arrive at your answer?

4) Water did not leak from the rectum while introducing water

**Comprehension:** What does water did not leak mean to you?

**Decision:** How hard is it to answer this question?

**Response:** How did you arrive at your answer?

5) My body position felt comfortable throughout irrigation

**Comprehension:** What does feeling comfortable mean to you?

**Decision:** How hard is it to answer this question?

**Response:** How did you arrive at your answer?

6) I experienced no pain during irrigation

**Comprehension:** What does pain during irrigation mean to you?

**Decision:** How hard is it to answer this question?

**Response:** How did you arrive at your answer?

7) I needed minimal hand strength to carry out my irrigation.

**Comprehension:** What does minimal hand strength mean to you?

**Decision:** How hard is it to answer this question?

**Response:** How did you arrive at your answer?

8) Using the irrigation product was straightforward.

**Comprehension:** What does straightforward mean to you?

**Decision:** How hard is it to answer this question?

**Response:** How did you arrive at your answer?

9) I was able to carry out irrigation without assistance from others

**Comprehension:** What does without assistance mean to you?

**Decision:** How hard is it to answer this question?

**Response:** How did you arrive at your answer?

10) The irrigation product was easy to take apart after use

**Comprehension:** What does easy to take apart mean to you?

**Decision:** How hard is it to answer this question?

**Response:** How did you arrive at your answer?

11) The irrigation product was easy to clean after use

**Comprehension:** What does easy to clean mean to you?

**Decision:** How hard is it to answer this question?

**Response:** How did you arrive at your answer?

12) My hand was not soiled with poo during irrigation

**Comprehension:** What does hand soiled mean to you?

**Decision:** How hard is it to answer this question?

**Response:** How did you arrive at your answer?

13) I was able to use the irrigation product away from home

**Comprehension:** What does away from home mean to you?

**Decision:** How hard is it to answer this question?

**Response:** How did you arrive at your answer?

14) I felt like my bowel was emptied after the irrigation

**Comprehension:** What does feeling like your bowel has been emptied mean to you?

**Decision:** How hard is it to answer this question?

**Response:** How did you arrive at your answer?

15) Irrigation gave me a sense of control over my life and daily activities.

**Comprehension:** What does a sense of control mean to you?

**Decision:** How hard is it to answer this question?

**Response:** How did you arrive at your answer?

**Part Two: Recall**

When answering these questions, what type of strategies did you use to retrieve information?

Are you considering the worst day/ week/ episode within the month, or are you using an estimate or average?

**Part Three: Overall**

1. What did you think about the number of questions in the Bowel Irrigation Questionnaire?
2. What do you think about the formatting of the Bowel Irrigation Questionnaire?
3. Were there any questions you felt unwilling to answer?
4. What suggestions do you have for making the Bowel Irrigation Questionnaire easier to use?
