## Appendix 1 for "Development, content validation and reliability assessment of the Bowel Irrigation Questionnaire, a Patient-Reported Experience Measure for Transanal Irrigation: a multi-methods study"

**Appendix One. Interview Guide for Health Care Professionals**

**INTERVIEW GUIDE: Health Care Professionals**

**Understanding the irrigation experience**

**Target group: 9 diverse profiles with experience with rectal irrigation,** covering academia, health care professionals and users.

**Length of interviews:** 60 min.	**Internal Ipsos job reference:** 24-052267

| **Introduction** | **5 min** | to | **5 min** |
| --- | --- | --- | --- |
| **Respondent understanding** | **10 min** | to | **15 min** |
| **Getting to irrigation** | **20 min** | to | **35 min** |
| **Initiation & Training** | **10 min** | to | **45 min** |
| **Securing a good experience** | **10 min** | To | **55 min** |
| **Wrap-up** | **5 min** | to | **60 min** |

**2. RESPONDENT INTRODUCTION	(10 min / 15 min)**

***The purpose of this section is understanding the condition, journey and experiences of the respondent.***

2.1 [5 minutes] **Respondent introduction**

**Moderator introduces him-/herself** to begin with, covering the same aspects as we would like the respondent to cover: Occupation, experience and professional interests.

- To begin with, could you briefly tell me a bit about your work, workplace and department and the set-up related to irrigation?
  - Do you have a dedicated team/clinic to manage irrigation?
  - What is your role and responsibilities when it comes to irrigation?
  - Which type of irrigation devices do you have at your department?
    - High/low volume? Brands?
    - What do you think about the devices you have available? Does it cover your needs, or do you miss anything?
- Who refers irrigation patients to your department?
  - What is typically written in the referral for potential irrigation patients?
  - Which types of patients get referred?
  - How many patients are referred for potential irrigation?
  - How many ends up getting irrigation? What do you think about this share?
  - How often do you recommend irrigation to people struggling with bowel issues?
  2.2 [5 minutes] **Irrigation experience**
- What is the first thing that come to mind now that we discuss “rectal irrigation”?
- What is your perception of the procedure?
  - How do your peers and colleagues talk about the procedure?
    - What questions do your colleagues ask about irrigation?
    - Are any of your peers/colleagues hesitant about the treatment? Why?
  - Where in the treatment lines do you see irrigation, now and in the future?
    - Why do you think irrigation is not more broadly used?
    - Why do you think it is mostly a treatment administered by specialists?
- Can you walk me through the first time that you became aware of irrigation as a possible treatment option for your patients?
  - How, when, why, by who were you introduced to irrigation?
  - What was your initial reaction to the treatment?
- What training or education have you been offered to learn about irrigation?
  - How did you experience these offers?
- What is your current view an s a treatment option to manage bowel issues?

**3. GETTING TO AN IRRIGATION DEVICE	(20 min / 35 min)**

***The purpose of this section is understanding thoughts, considerations and routes to an irrigation device.***

3.1 [4 minutes] **Case: Latest recommendation of irrigation¨**

**MODERATOR NOTE:** Further, the focus here is to understand pros and cons for irrigation in comparison to other treatments.

Try to remember the last time you recommended irrigation to a patient.

- Tell me about the patient and their situation: diagnoses, age, severity, anamnesis etc.
- How did you end-up suggesting irrigation? What steps lead to the decision?
  - How did the patient get assessed, and were there any other HCPs or departments involved in the treatment option?
  - What other treatments had been applied before irrigation?
- Why did you end up recommending irrigation for this person?
  - What thoughts and considerations lead to this decision?
  - What parameters influenced the decision?
  - What other treatment options were considered, if any?

**MODERATOR NOTE:** Treatment recommendations might not be relevant to all HCPs. In this case, please adapt to questions to the last patient you met using irrigation.

3.2 [4 minutes] **The route to irrigation**
- Can you describe the typical irrigation user (in your department)?
- How far into their “journey” do you typically see these patients?
  - What have happened before they see you?
  - What treatments have been administered?
- How far into their “journey” are patients typically prescribed irrigation?
  - Is this fitting – or would it ideally be prescribed earlier or later in the process?
- Why do you generally recommend irrigation?
  - What kind of symptoms are important to pay attention to?
  - What are the key factors you consider when recommending irrigation?
  - How does irrigation impact the lives of users? Positively and negatively?
    - What stories do you hear from patients?
- What are patients’ initial reactions to the irrigation?
  - What are common concerns and questions? How do you handle these concerns and questions? Can you provide any examples?
  - How much time are patients’ given to consider the treatment? What’s the process?
  - How much time typical pass from irrigation is first introduced until the patient’s start using the device? And why do you think that is?
  3.3 [4 minutes] **Choosing an irrigation device**
- What makes a good irrigation device in your experience? [Probe]
- Do you have one or more irrigation devices that you typically recommend? Why?
  - What do you like about it/them?
  - What sets it/them apart from competitors?
  - What can be improved on this/these devices?
- What is the best device that you have seen on the market?
  - What makes this product better than others?
- Are there any irrigation devices that you avoid? Why?
  - What makes this/these devices a worse option?
- If you have to sum up the characteristics of “a good device”, what are the five most important things?
  3.4 [4 minutes] **Patient involvement & preferences**
- How involved are the patients in the device decision? In what way?
  - What do the patients ask for?
  - What questions do they ask?
  - Do you see any typical preferences among your patients?
- Based on the feedback you have heard. What features, elements and components are irrigation users positively surprised by?
- What features, elements and components are irrigation users negatively surprised by?
- How important is the device design to patient? And what matters to patients when it comes to the design? (e.g. size, easy storing, discretion, softness etc.).
- What differences do you see across the different types of irrigation patients (e.g. neurogenic vs. functional, incontinence vs. constipation, age, gender etc.)
  3.5 [4 minutes] **Ideal product**

Let’s imagine you were commissioned to **design the ideal irrigation device** taking into account all your professional experience and the experiences shared by your patients. How would you go about it?

- What considerations are key to build the ideal product?
- What features would you prioritize?
- If you have to complete these sentences [encourage respondent to say multiple word at the end of each sentence]:
  - The device should be…
  - The device should make users feel…
  - The device should enable users to…
  - The device should definitely not…

**4. INITIATION & TRAINING	(10 min / 45 min)**

***The purpose of to understand the initiation and training phase from an HCP perspective.***

4.1 [10 minutes] **Introducing irrigation to a patient**

Imagine that you are sitting in front of a patient who you believe would benefit from irrigation.

- How would you introduce the treatment to a patient? Is there any adaptation according to the specific patient type?
- From your perspective, what kind of training do you believe is most beneficial for the patient?
- What training are the patients typically offered?
  - Do you offer home visits for initiation? Why/why not?
  - How do patients usually react to the training options available?
  - What options do they usually pick over the other?
  - What are the typical questions that patients would ask in relation to the training?
  - Are there any common concerns from patients?
- Do you offer your patients a product demonstration? Why / why not?
- To what extent do patients practically participate in a demonstrating?
  - Do you ask them to assemble/dissemble the device? Why/why not?
  - Do you ask the to perform an irrigation? On themselves or on a model? With/without the HCP in the room? Why/why not?
  - Do you use any sort of material to support your demonstration? (e.g. a rectum model, an anatomical model, drawings)? Why/Why not?
- Do you use any introduction support material, such as videos, images, brochures when introducing the device?
  - What material would you like to have available if you could choose freely?
  - What kind of training material works the best in your experience? (e.g. video/animations? Brochures? Web-based content? Rectum models?)
  - What defines good support material in your opinion?
- Are there any common difficulties patients face during the training phase?
- How are these addressed during and the session?
- How do you ensure patients know where to go for further questions or doubts after a session?
- What’s your perception of the training offered. How much is sufficient? How can it be improved?
- How important would you say training is overall in terms of ensuring a good experience with irrigation?

**5. SECURING A GOOD USER EXPERIENCE	(10 min / 55 min)**

***The purpose of to understand and define a good irrigation experience for user from a HCP perspective.***

5.1 [6 minutes] **Defining a good user experience**
- What does a "good irrigation experience" mean to you, both from a clinical and patient perspective?
- What makes a device user-friendly?
- When would you say that a patient has achieved a “good/successful irrigation experience?”
  - What are the most important attributes of an irrigation product that contribute to a positive experience? (e.g., ease of use, effectiveness, safety, patient comfort)
  - How does the irrigation products you currently use perform in terms of these attributes?
  - What parameters do you look at to measure “effectiveness” of irrigation for your patients?
- How important are usage instructions securing that patients have a successful irrigation?
- With everything that is currently available of educational material, support groups, product information…
  - How effective are these in supporting a good irrigation experience?
  - Are there any improvements or additions you would suggest?
- If you were to be responsible for improving patients experiences on irrigation, what would you do?
  5.2 [4 minutes] **Ensuring a positive irrigation experience over time**
- What are the key factors in ensuring that patients continue to use irrigation over time?
- What are the potential barriers to patient adoption or adherence to irrigation?
  - How do you address these challenges and provide support?
  - What support is offered to regular users after initiation?
    - Are home visits offered after initiation? Why /why not?
    - What do you think about it – is it sufficient? Why/why not?
- Do you have an example of a patient who dropped out of the treatment? What happened? When in the journey did the patient drop out?

**5. WRAP-UP	(5 MIN / 60 MIN)**

**In this section we wrap up the conversation and follow-up on any unanswered questions.**

- *Ask questions from client/backroom if any.*
- Is there anything that is on your heart or mind that we have not covered today?
- Do you have any questions for us before we wrap-up?

**Thank and close the interview.**
